# Pretreatment Attrition, Treatment Delay and Treatment Outcome in Drug-Resistant Tuberculosis: A Mixed-Methods Cohort Study in Public and Private Hospitals in Indonesia

**DOI:** 10.64898/2026.09.29.26364370

**Authors:** Farchan Azzumar, Aulia Fitri Rhamadianti, Eghar Anugrapaksi, Muhammad Prasetio Wardoyo, Assica Permata Amalya Hakiman, Luthfiyah Zanida Putri, Neni Sawitri, Kemalasari, Riyadi Sutarto, Ahmad Fuady, Erlina Burhan

**Author notes:** Correspondence: Prof. Erlina Burhan.

## Abstract

**Background:** Drug-resistant tuberculosis (DR-TB) remains a major problem in Indonesia, with a significant low treatment initiation rate. This study identifies factors influencing treatment initiation and assess the impact of delay on patient outcomes.

**Methods:** We conducted a mixed-methods study using a parallel convergent design. The quantitative component analyzed the Tuberculosis Information System (TBIS) data from three hospitals in Jakarta and Bogor from January 2022 to December 2023. The qualitative component involved in-depth interviews and Focus Group Discussions (FGD) with DR-TB patients, healthcare workers, patient supporters, and program managers.

**Results:** Among 1,665 people with DR-TB included in this study, we found a pre-treatment attrition rate of 6.4%. We identified two factors of pre-treatment attrition: prior TB treatment (aOR=1.7, 95%CI 1.082–2.671) and distance to hospital (aOR=1.9, 95%CI: 1.136–3.180). Conversely, referred patients, treatment in suburban areas, and people with rifampicin-resistant (RR)-TB were 85%, 82%, and 63% less likely to experience attrition, respectively. Qualitative findings identify patient, social, and health system barriers to the treatment initiation, and alternative remedies emerged as a coping mechanism.

**Conclusion:** Strengthening referral systems, decentralizing care, and addressing socio- economic barriers are critical for ensuring timely DR-TB treatment initiation.

## INTRODUCTION

Drug-resistant tuberculosis (DR-TB) remains a critical threat to global health with an estimated 400,000 new cases of multidrug- or rifampicin-resistant TB (MDR/RR-TB) in 2023 (1). Indonesia, a country designated by World Health Organization (WHO) as one of tuberculosis (TB) high-burden countries, bears an estimated 1.09 million new TB cases in Indonesia, including nearly 30,000 drug-resistant tuberculosis (DR-TB) new cases in 2023 (1). However, only 12,023 (41%) of DR-TB cases were diagnosed, and merely 8,259 (69%) of DR-TB notified cases initiated treatment in 2023 (2). Both numbers represent the magnitude of undiagnosed and untreated DR-TB cases that potentially transmit infection and undermine the efforts towards TB elimination.

The introduction of Xpert MTB/RIF and later Xpert Ultra has significantly improved the accuracy and turnaround time of diagnosing DR-TB. According to the recent systematic review that includes 86 studies in 95 countries, both Xpert MTB/RIF and Xpert Ultra yielded sensitivity of 95% [95%CI 91% to 98%] vs 95% [95%CI 90% to 98%] and specificity of 98% [96%CI 97% to 99%] vs 98% [95%CI 97% to 99%] for detecting rifampicin resistant (3). Despite these advances in diagnostic capacity, Indonesia continues to experience post-diagnostic attrition—a critical failure in the care pathway between diagnosis and treatment initiation, often termed pretreatment attrition (4).

Recent evidence highlights that pretreatment attrition is a pervasive and complex challenge in high-burden settings. A systematic review from six sub-Saharan African countries identified health system barriers—including laboratory operational delays, inadequate provider knowledge, and poor linkage between public and private sectors— as major drivers of delayed or missed treatment initiation (5). Similarly, a cascade-of- care analysis in urban Indonesia revealed that only 25.5% of diagnosed RR-TB patients successfully completed treatment, with substantial delays and losses concentrated between diagnosis and treatment initiation, particularly among those engaged with the private sector (6). In China, where pre-treatment attrition reached 36%, longer diagnosis delays were an independent predictor of pretreatment attrition, underscoring the time- sensitive nature of linkage to care (7). Together, these studies indicate that even with improved diagnostics, health systems struggle to translate diagnosis into timely treatment, and that patient pathways—especially those involving private healthcare— remain poorly understood.

Therefore, this mixed-methods study primarily aims to estimate the magnitude and determinants of pretreatment attrition and delays among DR-TB patients in Indonesia, and qualitatively explore the care-seeking pathways, health system bottlenecks, and patient experiences that explain these outcomes, with particular attention to differences between public and private hospital settings. In addition, this study also explores the DR-TB treatment outcomes between delayed and timely treatment initiation.

## METHODS

### Study design

We conducted a sequential mixed-methods study to investigate the post-diagnosis care cascade for Drug-Resistant Tuberculosis (DR-TB). The study was performed at three purposely selected Programmatic Management of Drug-Resistant Tuberculosis (PMDT) hospitals: Persahabatan Hospital Jakarta, Jakarta Islamic Hospital Cempaka Putih, and Dr. M. Goenawan Partowidigdo Pulmonary Hospital (RSPG Cisarua Bogor). These sites were selected to represent variation in institutional type (public and private) and settings (urban and suburban). This study enrolled people diagnosed with DR-TB between 1 January 2022 to 31 December 2023.

### Quantitative Component

This retrospective cohort was constructed from the National Tuberculosis Information System (TBIS). We have downloaded the TBIS data on 31 March 2024. We linked two keys register from TBIS, which are the TB06 form (list of individuals with presumptive TB, including those with confirmed DR-TB) and TB03 RO form (list of individuals initiating DR-TB treatment). To integrate both datasets, we performed a data merging procedure in SPSS. The TB03 RO dataset was merged with the TB06 dataset using unique patient identifiers. After resolving duplicate entries and inconsistencies, the final merged dataset consisted of 1,665 individuals. The cohort included only individuals in the final analyzable dataset aged ≥15 years with a bacteriologically-confirmed DR-TB diagnosis at three sites. We excluded individuals with pregnancy and incomplete data on diagnosis confirmation or key dates (diagnosis, treatment initiation), or missing essential demographic or clinical variables.

We collected information on sociodemographic and clinical characteristics, i.e., age, sex, HIV and diabetes status, DR-TB classification, previous TB diagnosis, hospital type and location, referral status, treatment initiation time, and treatment outcomes. Treatment outcome defined as treatment failures and mortality. Distance between the patient’s residence and the hospital was measured using Quantum Geographic Information System® (QGIS) version 3.34. The distance were then classified as ‘close’ (≤7.3 km) and ‘far’ (>7.3 km) (8).

In this study, pre-treatment attrition was defined as the failure to initiate DR-TB treatment within six months of diagnosis. Although there is no standard cut-off of pre-treatment attrition, the six-month threshold was chosen based on previous studies (9,10). In addition, treatment delay was defined as treatment initiation occurring more than seven days after diagnosis (11).

### Qualitative data collection

In the qualitative approach, we interviewed people with DR-TB who experienced pre-treatment attrition at the three hospitals. Participants were selected purposely, considering factors such as age, sex, and healthcare facility. Participants were contacted through telephone numbers listed in the TBIS and patient supporter networks. Qualitative data collection started from December 15th 2023 until January 15th 2024. In-depth interviews were conducted online or in person using structured interview guides in Bahasa Indonesia and were recorded. Each session lasted between 20 and 60 minutes and continued until data saturation was reached.

Additionally, we also did five focus group discussions (FGD) to explore factors affecting pre-treatment attrition and treatment completion. We invited representatives of physicians, nurses, patient supporters, and program managers at three hospitals into four separated FGDs according to professions. Another FGD, consisting of physicians and nurses from three primary health cares, Puskesmas Pulogadung, Puskesmas Cempaka Putih, Puskesmas Cisarua, were conducted separately. A single data coder transcribed and processed the interviews without returning transcripts to respondents. All interviews were conducted in Bahasa Indonesia and audio-recorded with participants’ consent.

### Data management and statistical analysis

For quantitative analysis, datasets from both TB03 and TB06 forms in the three hospitals were merged and stored in an MS Excel database. Data cleaning and validation were applied before analysis—all using STATA version 17.0 software. We have access to each participant identifying information during the data collection process and anonymizing the data during the analysis process. Descriptive analyses summarized sociodemographic characteristics, treatment status, and clinical profiles of DR-TB cases. Bivariate and multivariate logistic regression analyses were performed to assess factors associated with treatment initiation (whether the patient started treatment) and treatment delay (whether the patient experienced a delay in treatment). The association between the interval from diagnosis to treatment initiation and patient outcomes was examined using Cox regression analysis. Each independent variable was first analyzed individually using the enter method, with non-significant variables sequentially eliminated. A variable was considered to have a significant influence on the outcome if the p-value was less than 0.05.

For qualitative, interview transcripts were transcribed verbatim, verified for accuracy, and analyzed by inductive thematic analysis to identify emerging themes (12). Key terms were systematically coded in Microsoft Excel, incorporating both data-driven and literature-based themes. Interviews and focus group discussions were independently coded, with emerging themes validated and categorized as barriers or facilitators to DR-TB treatment initiation (13).

### Ethical approval

The study received ethical approval from the Ethics Committee of the National Respiratory Center, RSUP Persahabatan (Approval number of original ethical clearance 172/KEPK-RSUPP/12/2023, dated December 15, 2023 and the amendment protocol 0172.A.1/KEPK-RSUPP/12/2024, dated December 27, 2024). As the quantitative part of this study involved retrospective use of existing data and did not involve direct contact with patients, individual informed consent was waived by the relevant ethics committee. For the qualitative sub-study, given that the study involved the analysis of secondary data, ethical approval was obtained for the use of questionnaires in the qualitative sub-study to ensure written or recorded verbal informed consent from participants. All procedures were conducted in accordance with ethical guidelines to protect participant confidentiality and uphold research integrity.

## RESULTS

### Clinical and sociodemographic characteristics

A total of 1,655 people with DR-TB were included in this study, of whom 1,559 (93.6%) initiated treatment, while 106 (6.4%) did not commence DR-TB therapy (Figure 1). The overall median interval from diagnosis to treatment initiation was 13 days (IQR: 8–24 days), with a mean duration of 22 days. The majority (95.4%) were aged between 15 and 65 years, while 4.6% were older than 65 years, with the mean age of the study population was 40.79 ± 14.65 years (Table 1). Previous TB history was comparable, with 49.2% (n = 819) of newly diagnosed cases and 50.8% had previously received TB treatment. Among patients with previous TB treatment 468 (28.1%) were relapse, 165 (9.9%) cases of treatment default, and 213 (12.8%) treatment failure cases. Rifampicin-resistant TB (RR-TB) was the most common classification (51.2%, n = 853), followed by multidrug-resistant TB (MDR-TB) (37.7%), isoniazid-resistant TB (Hr-TB) (19.7%), pre-XDR-TB (8.3%), XDR-TB (0.8%), and poly-resistant TB (0.1%). Most people with DR-TB received treatment at public hospitals, with 41.6% (n = 692) at RSUP Persahabatan and 43.9% (n = 731) at RSPG Cisarua. Additionally, 81.0% of patients were referred from other healthcare facilities before initiating DR-TB treatment.

**Figure 1.**
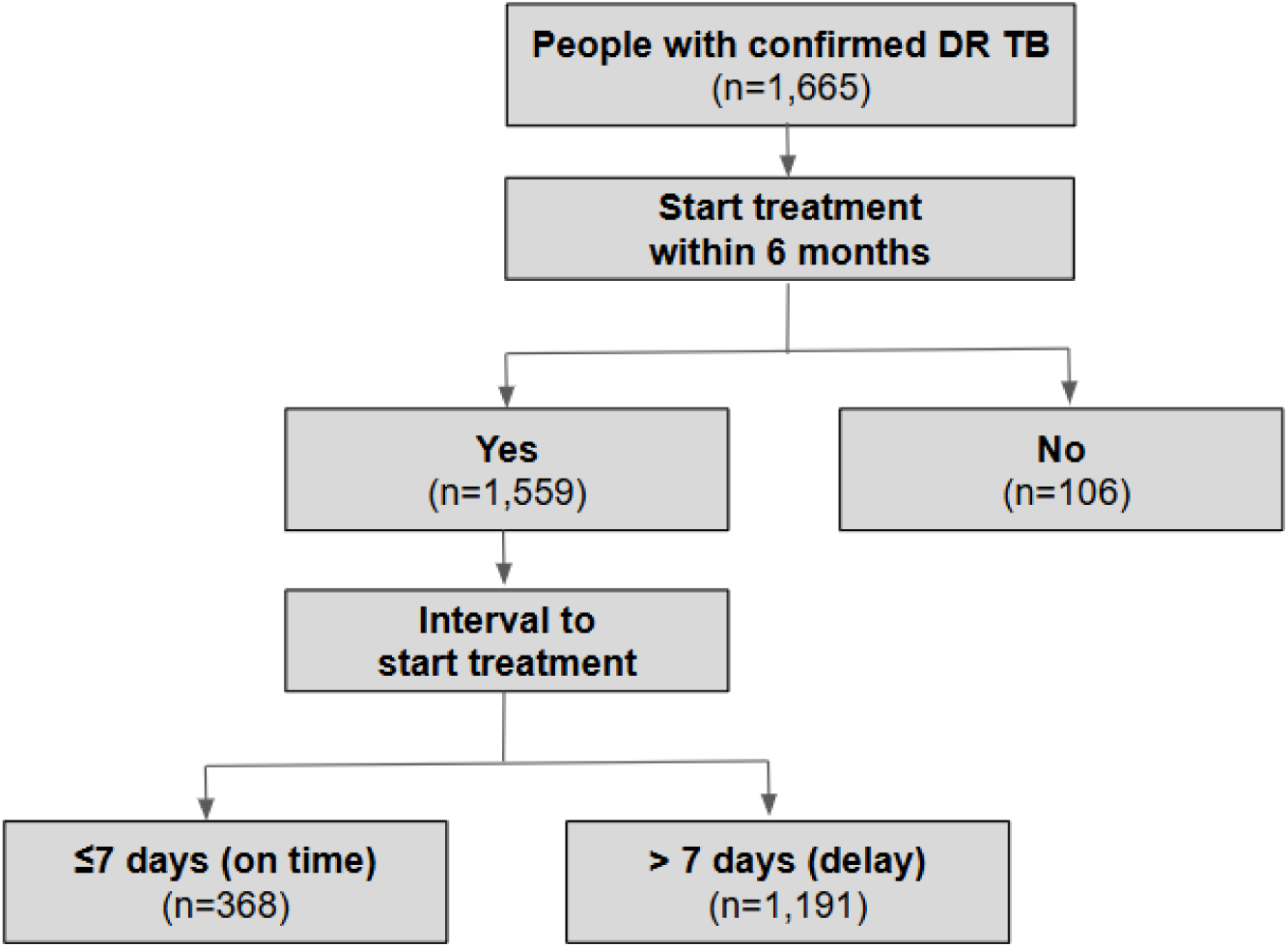
Study sample flowchart.

**Table 1.**
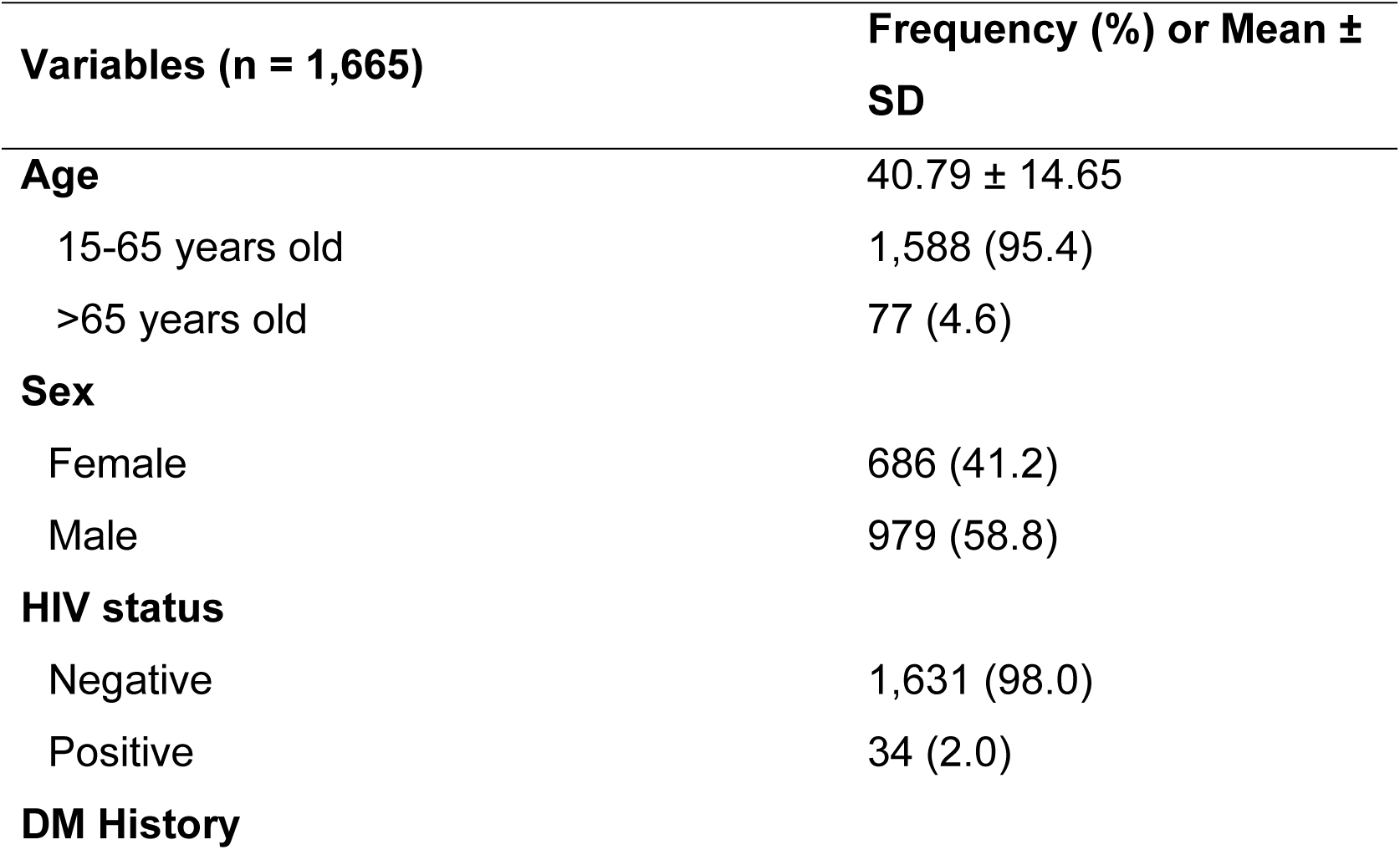
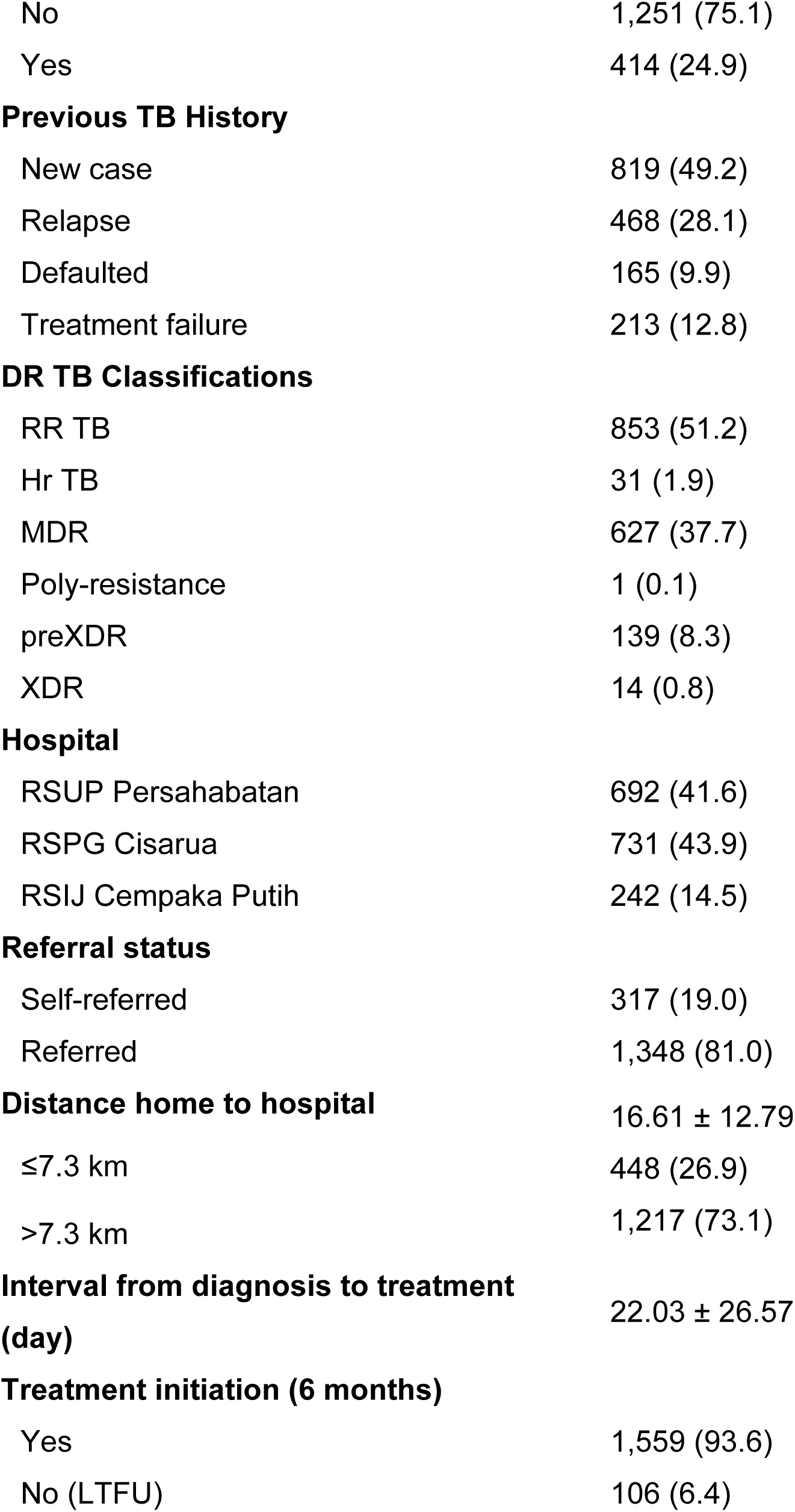
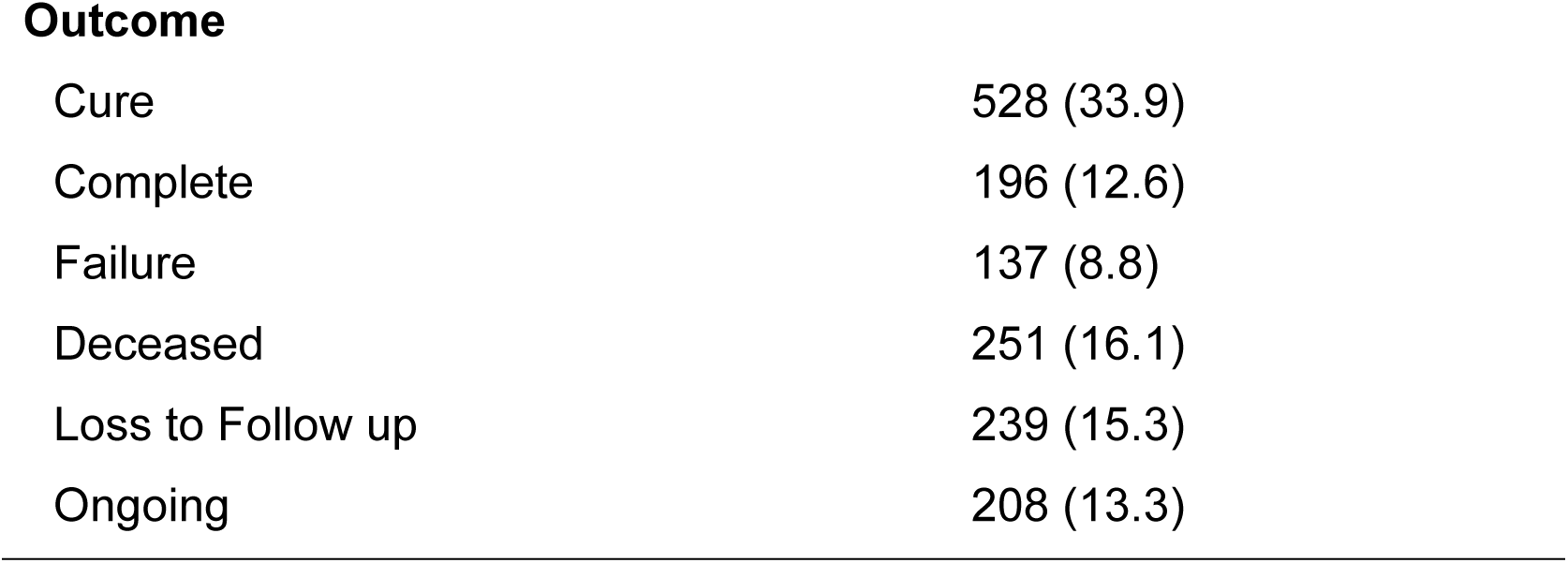
Clinical and socio-demographic characteristics of people with DR-TB at three designated hospitals.

| Variables (n = 1,665) | Frequency (%) or Mean ± SD |
| --- | --- |
| **Age** | 40.79 ± 14.65 |
| 15-65 years old | 1,588 (95.4) |
| >65 years old | 77 (4.6) |
| **Sex** |  |
| Female | 686 (41.2) |
| Male | 979 (58.8) |
| **HIV status** |  |
| Negative | 1,631 (98.0) |
| Positive | 34 (2.0) |
| **DM History** |  |
| No | 1,251 (75.1) |
| Yes | 414 (24.9) |
| **Previous TB History** |  |
| New case | 819 (49.2) |
| Relapse | 468 (28.1) |
| Defaulted | 165 (9.9) |
| Treatment failure | 213 (12.8) |
| **DR TB Classifications** |  |
| RR TB | 853 (51.2) |
| Hr TB | 31 (1.9) |
| MDR | 627 (37.7) |
| Poly-resistance | 1 (0.1) |
| preXDR | 139 (8.3) |
| XDR | 14 (0.8) |
| **Hospital** |  |
| RSUP Persahabatan | 692 (41.6) |
| RSPG Cisarua | 731 (43.9) |
| RSIJ Cempaka Putih | 242 (14.5) |
| **Referral status** |  |
| Self-referred | 317 (19.0) |
| Referred | 1,348 (81.0) |
| **Distance home to hospital** |  |
|  | 16.61 ± 12.79 |
| ≤7.3 km | 448 (26.9) |
| >7.3 km | 1,217 (73.1) |
| **Interval from diagnosis to treatment (day)** |  |
|  | 22.03 ± 26.57 |
| **Treatment initiation (6 months)** |  |
| Yes | 1,559 (93.6) |
| No (LTFU) | 106 (6.4) |
| Outcome |  |
| Cure | 528 (33.9) |
| Complete | 196 (12.6) |
| Failure | 137 (8.8) |
| Deceased | 251 (16.1) |
| Loss to Follow up | 239 (15.3) |
| Ongoing | 208 (13.3) |

### Factors associated DR-TB pre-treatment attrition

In the multivariate analyses, pre-treatment attrition was associated with previous TB history, DR-TB classifications, hospital location, referral status, and distance to hospital. Individuals with previous TB treatment history had a 1.7-fold higher likelihood of not initiating treatment compared to newly diagnosed DR-TB (aOR = 1.7, 95% CI: 1.082– 2.671). Notably, after adjusting for confounders, patients residing >7.3 km from the treatment facility exhibited a 1.9-fold increased risk of pre-treatment attrition compared to those living closer, suggesting that greater distance is associated with lower treatment uptake. Conversely, patients classified as rifampicin-resistant, those receiving treatment in suburban hospitals, and those referred by healthcare providers had significantly lower odds of missing treatment, with reductions of 63%, 82%, and 85%, respectively.

### Factors associated with DR-TB delayed treatment

Among 1,559 patients initiating treatment, only 24% started the treatment within the first seven days, while the majority (76%) experienced a delay >7 days after diagnosis. We found that female patients, those with prior TB history, referred cases, and those treated in urban or public hospitals had a higher odds of treatment delays. Males had significantly lower odds of treatment delays compared to females (aOR = 0.73, 95% CI: 0.558–0.947), (Supp. Table 1). Additionally, those treated in private or suburban hospitals had a significantly reduced likelihood of delayed treatment, by 91% and 82%, respectively. Interestingly, although referred patients were more likely to initiate treatment, they had a 3.9-fold higher probability of experiencing treatment delays compared to self-referred patients (aOR = 3.85; 95% CI: 2.806–5.291). Individuals with previous TB treatment also had a 1.32-fold higher likelihood of experiencing delays, indicating that previously treated patients are at greater risk for both pre-treatment attrition and delayed treatment initiation.

### Delayed treatment initiation correlates with unfavorable outcomes

Patients with ongoing treatment were excluded due to unavailable outcomes, yielding a final sample of 1,351. Among them, 528 (39%) were cured, 196 (14%) completed treatment, 251 (19%) failed treatment, 137 (10%) died, and 242 (18%) were lost to follow-up. Figure 2 illustrates that treatment delays exceeding seven days significantly increase the risk of treatment failure. As shown in Table 3, delayed initiation was associated with a higher hazard of treatment failure (HR = 1.83; 95% CI: 1.127– 2.990). Moreover, prolonged time to treatment was associated with delayed *Mycobacterium tuberculosis* culture conversion at Months 3, 4, 5, and 6, with the highest odds observed at Month 6 (OR = 1.02, 95% CI: 1.006–1.023), though the risk increase was marginal (Supp. Figure 1 and Supp. Table 2).

**Figure 2.**
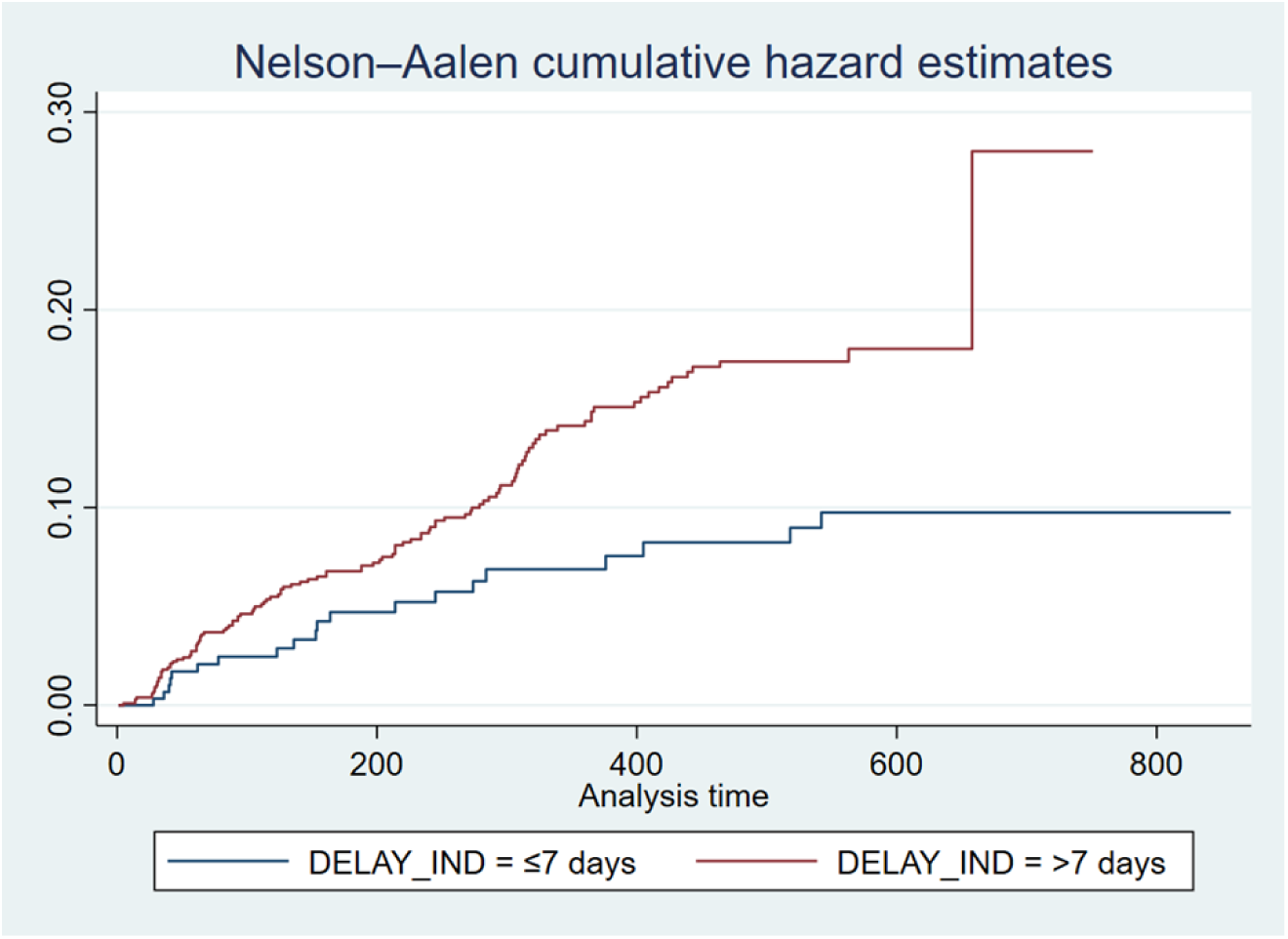
Nelson-Aalen Cumulative Hazard Curve for Treatment Failure.

**Table 2.**
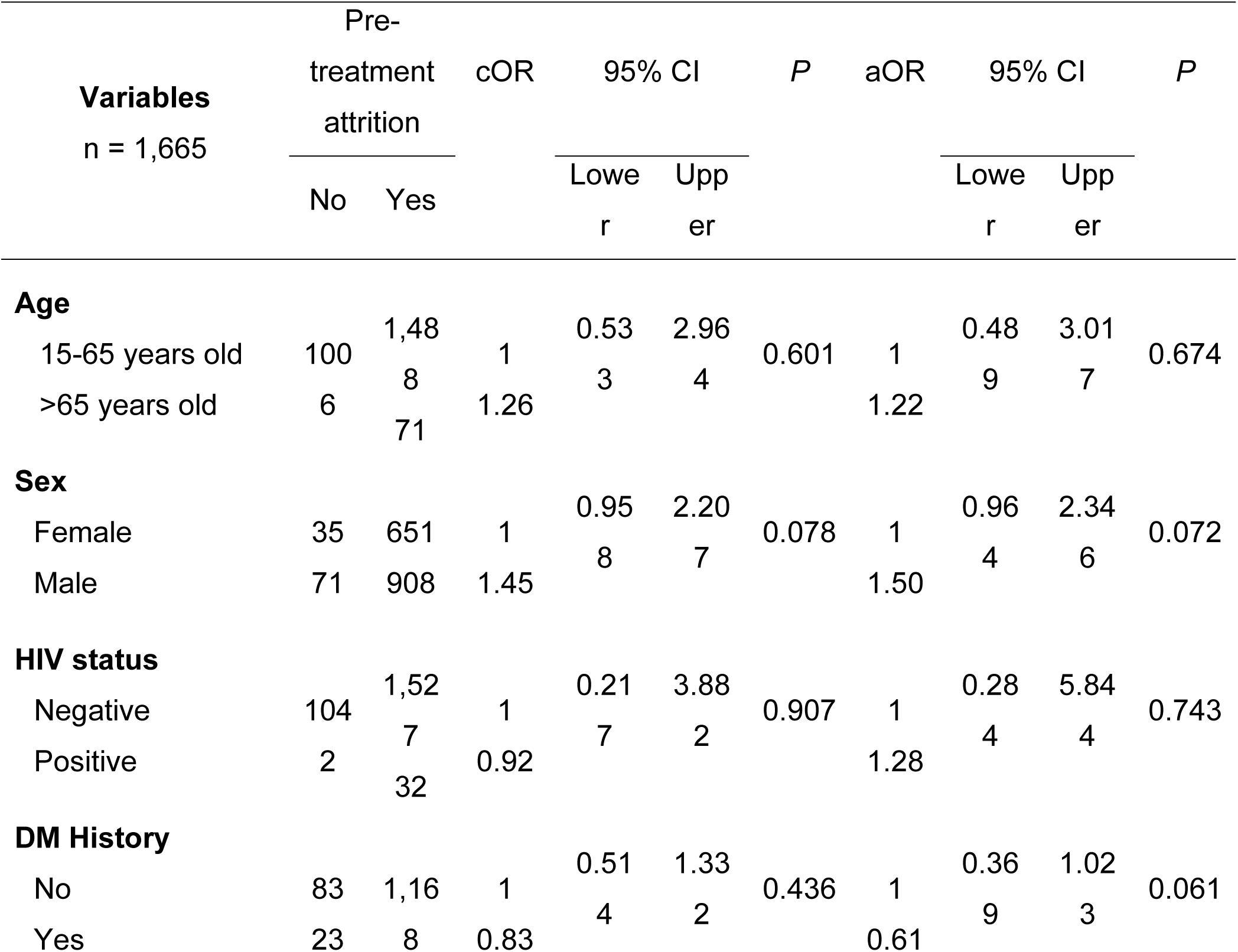
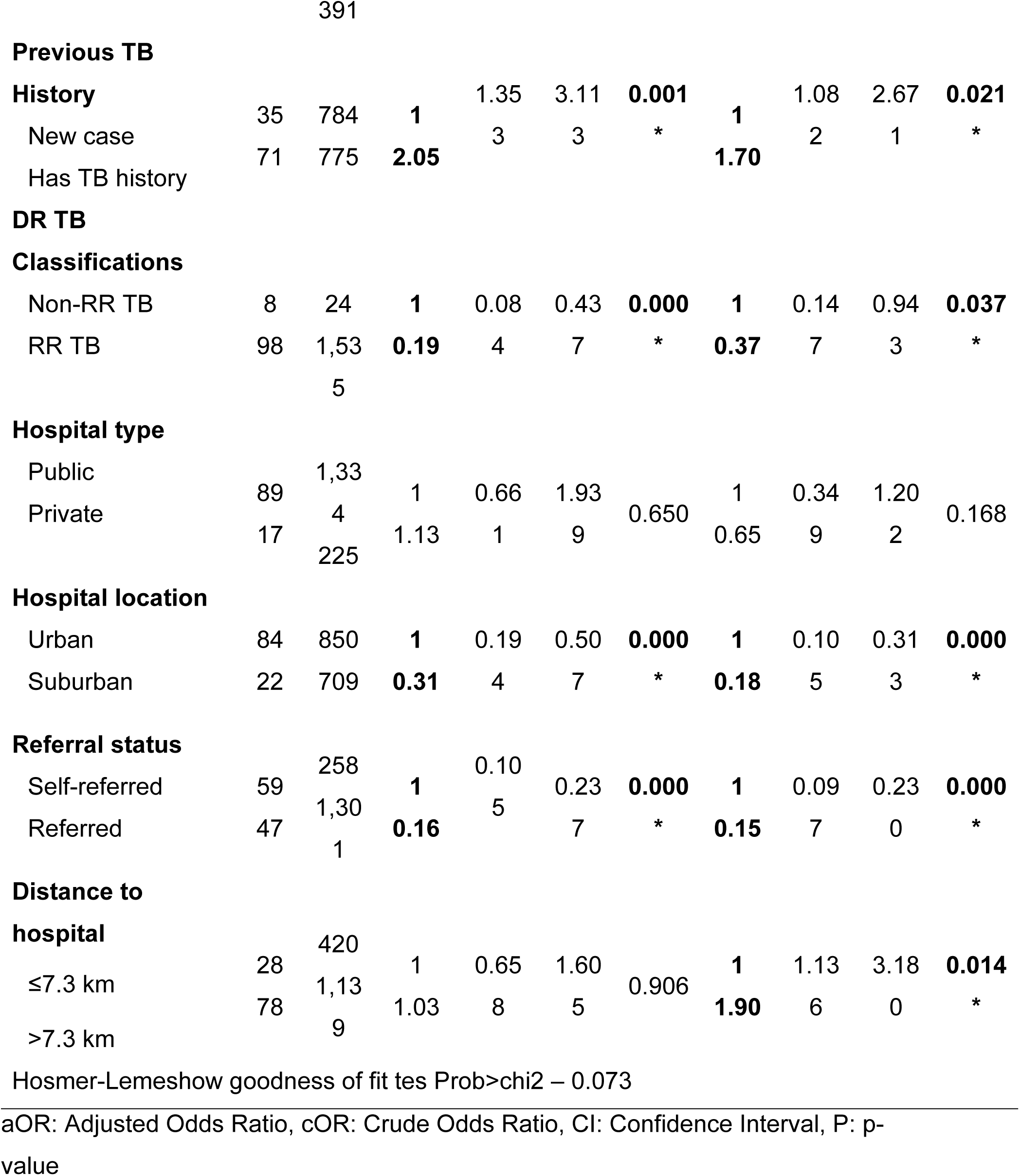
Univariable and Multivariable Logistic Regression of Factors Associated with pre-treatment Attrition.

**Table 3.** Multivariate Cox Regression Analysis of Treatment Failure in Patients with Drug-Resistant Tuberculosis.

| Variables<br>n = 1,351 | Outcome |  | HR | 95% CI |  | P |
| --- | --- | --- | --- | --- | --- | --- |
|  | Failure<br>(event) | Other<br>outcomes<br>(censored) |  | Lower | Upper |  |
| Interval to begin treatment |  |  |  |  |  |  |
| ≤7 days | 20 | 288 | 1 | 1.12 | 2.99 | 0.015 |
| >7 days | 117 | 926 | 1.83 | 7 | 0 | * |
| Age |  |  |  |  |  |  |
| 15-65 years old | 133 | 1,154 | 1 | 0.32 | 2.36 | 0.784 |
| >65 years old | 4 | 60 | 0.87 | 0 | 1 |  |
| Sex |  |  |  |  |  |  |
| Female | 67 | 513 | 1 | 0.57 | 1.13 | 0.226 |
| Male | 70 | 701 | 0.81 | 9 | 7 |  |
| Previous TB History |  |  |  |  |  |  |
| New case | 70 | 611 | 1 | 0.70 | 1.39 | 0.970 |
| Has TB history | 67 | 603 | 0.99 | 6 | 6 |  |
| Referral status |  |  |  |  |  |  |
| Self-referred | 20 | 200 | 1 | 2.54 | 1.44 | 0.627 |
| Referred | 117 | 1,014 | 0.88 | 1 | 7 |  |
| Distance to hospital |  |  |  |  |  |  |
| ≤7.3 km | 32 | 312 | 1 | 0.78 | 1.75 | 0.432 |
| >7.3 km | 105 | 902 | 1.17 | 7 | 0 |  |

HR: Hazard Ratio, CI: Confidence Interval, P: p-value

However, treatment delays were not significantly associated with mortality. Mortality was associated significantly with elderly individuals than younger individuals (HR= 3.49; 95% CI: 2.371-5.134), distance >7.3 km (HR= 1.46; 95% CI: 1.068-2.009) and self-referred (HR= 1.43; 95% CI: 1.038-1.975) (Supp. Table 3). These findings underscore the importance of timely treatment in mitigating treatment failure and ongoing transmission while highlighting key demographic and geographic risk factors for mortality in people with DR-TB.

### Qualitative data

From the dataset of confirmed DR-TB treatment statuses, in-depth interviews were conducted with 14 people with DR-TB or their caregivers. Additionally, one informant from East Jakarta, within the coverage area of Persahabatan Central General Hospital, was interviewed despite not appearing in the TBIS records. In total, 15 individual interviews but only 4 people qualified as informants (Supp. Table 4) and 31 informants across five Focus Group Discussions (FGD) (Supp. Table 5) were included. Our analysis identified key themes shaping each stage of the care cascade, categorized into individual-level factors, social, and health system factors. These findings provide insight into barriers and facilitators affecting timely treatment initiation and adherence.

### Individual Factors

#### a. Tuberculosis Stigma

We identified several factors affecting pre-treatment attrition, denial of being diagnosed with TB, lack of understanding regarding DR-TB, and lack of patient determination to seek treatment.

Denial of diagnosis poses challenges to treatment initiation, especially in DR TB cases. Denial was indicated when individuals preferred to be diagnosed with (residual)

stroke than TB, and denied having typical TB symptoms as "*the symptoms of TB are not very clear*," "*just a normal cough*," "*the symptoms are not very significant*," or "*feeling still healthy*." (Doctor 3).

This was supported by patient supporters, citing that "*sometimes, those who receive the treatment still haven’t accepted it.*" (Patient Supporter 1)

Denial often stems from limited knowledge about TB, DR-TB risks, and treatment management. Without comprehensive information, patients are more likely to reject their diagnosis. The denial may be associated with stigma perceiving TB as "*a disease of the poor*" or "*can lead to being separated from the family once diagnosed with drug-resistant TB*" (Patient Supporter 1).

Leading to shame and social isolation due to lack of understanding of TB in society. In addition to the fact that stigma can evoke negative feelings within the patient, it can also prevent DR-TB patients from receiving support from their families to seek treatment.

"*Some patients are ostracized and end up in shelters. Others are too ashamed to continue treatment.*" (Patient Supporter 1). This situation can arise due to stigma in the workplace, where companies are considered reluctant to allow sick leave for people with DR-TB patients to seek treatment. "*But it’s actually not the patient’s obstacle, but rather it’s because of constraints in the office, which are somewhat inflexible in allowing their employees to take repeated leave for treatment, forcing patients to choose between their job and their health.*" (Puskesmas Health Worker 1)

#### **b.** Concerns on long treatment and adverse effects

Treatment concerns are also a factor that inhibits patients from seeking treatment, due to the lack of understanding of patients regarding the safety of anti-tuberculosis medications, resulting in patients fear of the side effects and choosing alternative treatments as coping.

Informants expressed fear and concerns regarding DR-TB treatment’s adverse effects, the quantity of drugs, and the duration of treatment. Educational messages from healthcare workers and during home visits may trigger these concerns as doctors explained the side effects and number of medicines, but persons with DR-TB even worried.

*"The doctor explained the side effects (like) muscle weakness, nausea, stomach issues, possible hearing loss, hallucinations, and skin darkening. It’s overwhelming, especially taking 16 pills at once. Won’t it harm other organs?"* (Patient 4), then they decided to not undergo the treatment because of that worry.

Healthcare workers acknowledge these fears, noting that some patients refuse treatment despite home visits and education. From the patient’s perspective, learning about the side effects often intensifies their anxiety, making treatment initiation even more difficult.

### Social Factors

#### a. Family Support

Family plays a crucial role in TB treatment adherence. Patients with strong family support are more likely to start and continue treatment, while those without it face significant challenges. This sentiment is expressed by doctors, nurses, and patient supporters, all agreeing that the role of family support is vital to help patients start treatment.

Some respondents noted that family support is crucial because people with TB “definitely need family (support)” (Patient Supporter 1) and those with sufficient support “*tend to succeed (in completing treatment).*” (Patient 5)

#### **b.** Distance to healthcare facilities

Distance is a major barrier to treatment initiation, particularly for patients living far from healthcare centers. One family of a non-treating patient expressed that distance significantly affects treatment initiation which is confirmed by healthcare workers who directly handle people with DR-TB daily. Distance is one of the things that make patients reluctant to start treatment.

Doctors cited that people with treatment delay usually come from far distance from the facility, such as "*When we receive patients, they usually come from West Jakarta, it’s also far, (from East Jakarta)*" (Doctor 1).

#### **c.** Economic Burden

Economic constraints, including transportation expenses and loss of income, significantly impact patients’ willingness to start treatment. Some patients delay or refuse treatment because they must choose between their health and earning a living, especially for those who are the "*breadwinners*" (Patient Supporter 3). Patients believe that DR-TB treatment, including dealing with its side effects, takes time and potentially causes them or their supportive family members to lose income. It is confirmed by doctors based on their conversation with patients.

> *" It’s always an economic issue. If a patient stops working, their income decreases. Some say, ’I have no one to take me, Doc,’ or ’I have to work, Doc." (Doctor 6).*

#### **d.** Peer’s Experience factor

Experiences they hear from peers , such as “his body turned black, he couldn’t get up for three months, and his kidneys and liver were affected” (Patient 4) can increase worry and anxiety when not properly delivered. It also needs "They need repeated education and support from family or partners." (DR-TB Program Manager 1)

### Health System Factors

#### a. Lengthy process at Hospital

Multiple visits for baseline assessments and the complex procedures can lead to frustration and hindered treatment initiation. Clinical assessment which can be done within two days took *“longer time (and multiple visits) because they didn’t receive service on that day."* (Patient Supporter 3) Lack of facility to perform baseline assessment contributed to the lengthy process because “(patients need to be) *referred back to Persahabatan Central General Hospital.”* People with TB *may “feel like they’re being tossed around* (DR-TB Program Manager 5) and can lead to treatment delay and attrition. Another issue was quota–a maximum maximum number of patients can be served in one facility, usually 20 or 30 patients per day. This leads to unnecessary referral.

#### **b.** Healthcare Workers-Patient Communication

Unclear communication, including errors in delivering DR-TB diagnosis, such as “I was told it was negative. Instead, I was put on COPD treatment” (Patient 1) and “They just said they didn’t have TB. Just because there’s a lot of phlegm” (Patient 3), leads to treatment delay. This information was perceived lacking at the primary level care and instead only referred individuals with DR-TB to secondary hospitals without sufficient information. This led to confusion for those diagnosed and delay in treatment initiation.

> *“Patients need to understand why they are being referred and why they can’t be treated at the community health center." (DR-TB Program Manager 7).*

#### **c.** Unpleasant Treatment Experiences

Patients at DR-TB hospitals face unpleasant experiences for example long waits that were “tiring” and “exhausting” because they had to register at 7am but completed the queue at 1pm, then waited for the medicine (Patient 3 and Patient 4). Their failure in the previous treatment course “*even after over a year of treatment”* made them “tired” and “gave up” with the treatment. (Doctor 2).

#### **d.** Alternative Remedies as a Coping Method

Instead of starting treatment, because they “*don’t want to take risks (of adverse effects)” (Patient 4)*, individuals with DR-TB opted for alternative remedies to address the DR-TB because “*that doesn’t have side effects on other organs.”* (Patient 4). They feel safer with alternative remedies and easier to accept if something happened after receiving such remedies, citing “if it is fate (the death after receiving alternative remedies, *we leave it to The One Above (God)*" (Patient 4).

## DISCUSSION

This study reveals that the pre-treatment attrition of DR-TB in three hospital in Indonesia is low (6.4%) compared to WHO Global Report, Indonesia (17%). Even though this study result may not represent the nationwide population, this study propose to clearly define the time window of observation since the date of diagnosis.

This study examined factors influencing TB treatment initiation and delays using a mixed-method parallel convergent design. Our findings indicate that 94.2% of patients in our sample initiated DR-TB treatment, significantly higher than the national average of 68% in Indonesia, likely due to better healthcare accessibility. Factors facilitating treatment initiation included receiving care in suburban areas, being rifampicin-resistant groups, and being referred, with referred patients having an 85% lower likelihood of pre-treatment attrition. The lower attrition in the RR group may be attributed to the national diagnostic protocol, which utilizes GeneXpert testing for rifampicin resistance, leading to earlier treatment initiation (11). These findings underscore the influence of clinical characteristics and geographic barriers on treatment initiation, highlighting key areas for targeted interventions to enhance early treatment uptake.

Conversely, a history of DR-TB and long travel distances were associated with treatment delays. This aligns with the qualitative findings, revealing that patients living far from healthcare facilities were reluctant to start treatment, similar to Warumo et al., who reported that long travel distances increased the hazard of loss to follow-up (LTFU) by six times (14). Additionally, our qualitative data corroborate Oga-Omenka et al., who found that patients with prior TB history often experienced treatment fatigue, making them less willing to restart prolonged medication regimens.^15^ Addressing geographic and psychosocial barriers through strengthened referral systems may enhance DR-TB treatment uptake.

In further investigating factors contributing to treatment delays, our findings revealed that, unexpectedly, male patients were less likely to experience delays compared to female patients, despite potential occupational constraints. This aligns with previous studies indicating that female patients have a higher risk of delayed treatment initiation (15–17). Horton et al. suggested that this disparity may stem from the fact that tuberculosis is more prevalent among men in low- and middle-income countries, potentially leading to less attention being given to early treatment initiation for women (18). Additionally, treatment at private hospitals significantly reduced the likelihood of delays, with 91% of patients receiving treatment on time. Patients often navigated a complex healthcare pathway, seeking diagnoses and treatment from both private and public providers (19). However, waiting times were longer in public hospitals, while private facilities facilitated faster treatment initiation, highlighting their role in improving TB care access (20). Our findings also indicated that patients treated in urban hospitals were more likely to experience treatment delays. Although urban centers generally provide better healthcare access, they also encounter systemic inefficiencies, including overcrowded facilities, prolonged waiting times, and bureaucratic delays. These challenges were further emphasized in our qualitative findings, where patients expressed frustration over long queues, limited availability of healthcare staff, and quota-based treatment policies.

Our findings indicate that prior TB history and being a referred patient increased the likelihood of delayed treatment by 1.3 times and nearly 4 times, respectively. This aligns with previous results showing that individuals with prior TB treatment were often reluctant to restart therapy, leading to delays (21,22). Family support played a crucial role in motivating patients (13), as highlighted in our qualitative findings, where strong familial encouragement was associated with better treatment adherence. Although HIV increased the likelihood of delayed treatment by 1.7 times, the association was not statistically significant. Previous studies have reported similar findings, showing that individuals with a history of TB and those with severe illness, including HIV, are more likely to experience delays in treatment initiation (23). These findings underscore the need for targeted interventions to address barriers faced by high-risk groups, such as strengthening counseling services, enhancing family engagement, and streamlining referral systems to minimize delays.

We explored the impact of delayed treatment on patient outcomes, specifically treatment failure. Our qualitative findings highlighted that delays were often caused by the extensive pre-treatment assessment process. According to clinical guidelines, several mandatory evaluations—such as chest radiography, electrocardiography (ECG), complete blood count, and pregnancy testing—must be completed before treatment initiation (11). Notably, patients who initiated treatment more than seven days after diagnosis had a 1.8-fold higher risk of treatment failure, with prolonged culture conversion further supporting this finding. Previous studies have similarly reported that delayed treatment initiation is associated with poorer outcomes and extended culture conversion times (24,25). Regarding mortality risk, older age and long travel distances significantly increased the likelihood of death, consistent with prior research showing that advanced age, male sex, and HIV positivity were key mortality risk factors among patients receiving TB treatment (26). These findings underscore that delayed treatment reflects systemic inefficiencies, not only compromises outcomes but also prolongs infectiousness, reinforcing the need for timely intervention. Streamline diagnostic and pre-treatment procedures are crucial to prevent delayed treatment. Prolonged treatment initiation may contribute to poor adherence and disease severity, increasing the risk of treatment failure and delayed culture conversion.

Corroborating our quantitative findings, our qualitative analysis categorizes the barriers to DR-TB treatment initiation into three main domains: patient-related factors, healthcare system challenges, and social determinants.

### **a.** Patient Factors: Fear of DR-TB Treatment

The study highlights patients’ ignorance of DR-TB, concerns about OAT side effects, fears of stigma and discrimination, and perceptions of ineffective DR-TB services. Denial of DR-TB diagnosis may stem from patients feeling physically well, lacking classic TB symptoms, or attributing symptoms to other illnesses. This denial is exacerbated by patients’ low knowledge of DR-TB and fear of OAT side effects. Many patients struggle with lengthy and side effect- ridden DR-TB therapy regimens. However, information on OAT side effects should be provided cautiously, considering patient readiness. Counseling during diagnosis delivery could help address patient concerns and reluctance to seek treatment (5,27). Psychological counseling is also crucial for newly diagnosed people with DR-TB.

### **b.** Social Factors

Stigma, distance, family support, and economic conditions are recurring issues in DR-TB treatment. Negative perceptions toward people with DR-TB, leading to feelings of shame, hinder treatment initiation. Transportation is also a critical factor, yet distance remains a significant barrier. Efforts to decentralize DR-TB services are essential to improving accessibility. Economic burdens, including treatment costs and income loss, further hinder treatment initiation (27,28). In Indonesia, initiatives such as support community formation and service decentralization have been introduced to enhance access. Addressing stigma and geographic barriers requires integrated, cross-sectoral strategies and greater awareness of the psychosocioeconomic factors affecting treatment adherence. Institutionalized interventions are necessary to effectively identify and mitigate these challenges.

### **c.** Health System Factors

We emphasize the importance of patient-centered healthcare systems, focusing on efficient in-house services that minimize waiting times and visit frequencies. In some DR-TB referral hospitals, baseline examinations are ineffective due to limited schedules and insufficient human resources, particularly clinicians and data managers, essential for effective DR-TB services. Similar concerns are echoed in other studies, highlighting patient perceptions of DR-TB service effectiveness and issues with recording systems, data management, and notifications. Negative patient perceptions often stem from prolonged waiting times and coordination challenges between referring and referral healthcare facilities, notably in public healthcare settings (29). On the other hand, the risks of medication make patients choose alternative medicine over conventional healthcare. This is reasonable because according to the national survey results, 24.4% have used traditional healers or traditional medicine, and 32.9% have used it as the complement of conventional healthcare (30).

Both qualitative and quantitative findings indicate that prolonged waiting times in public healthcare facilities are associated with people with DR-TB’ reluctance to initiate or continue treatment. To address this issue, policymakers should focus on improving service quality, particularly by minimizing delays in treatment initiation. Effective communication is also a critical determinant for DR-TB treatment success. However, this study identifies significant gaps in conveying DR-TB diagnoses and coordination between healthcare facilities, which disrupt continuity of care. Strengthening communication strategies, including clarifying service procedures and improving coordination among providers, is essential to fostering patient trust and encouraging timely treatment initiation. As a suggestion to address unpleasant care experiences, decentralisation of health services should be implemented and if there are long queues, patient education can be provided to reduce patient burnout. Addressing these structural issues is crucial to optimizing DR-TB care delivery and improving patient outcomes.

A key limitation of this study is that the sample population was drawn from only three designated hospitals, which may not fully represent the broader healthcare landscape in Indonesia. This restricted scope could contribute to discrepancies in the treatment initiation rate observed in our study compared to national estimates. Future research with a more diverse and representative sample across various healthcare settings is needed to enhance the generalizability of these findings.

### CONCLUSIONS

This study highlights the critical role of timely DR-TB treatment initiation in improving patient outcomes and reducing treatment failure and delayed culture conversion. Prior TB history and geographical barriers contribute to treatment attrition and delays, while effective referral systems and decentralized care enhance access. The strong association between delayed treatment and poorer outcomes underscores the need to streamline diagnostic and pre-treatment procedures. Addressing multidimensional factors (patient awareness, social stigma, and healthcare system inefficiencies) is essential to optimizing the TB care cascade. Strengthening healthcare services, improving patient counseling, and mitigating socio-economic barriers are key to enhancing treatment adherence. Future research should incorporate nationwide data to validate these findings and assess the broader impact of proposed interventions.

### DECLARATION OF INTERESTS

The authors declare no conflict of interest.

### FUNDING

This study received a research funding grant from the PENABULU-STPI Consortium Community as an external sponsor. The sponsor had no involvement in conducting the study.

### AUTHOR CONTRIBUTIONS

FA, EA, RS, EB, and AF contributed to the conceptualization and methodology of the study. Project administration was carried out by FA, EA, MPW, and APAH, with EB providing supervision and validation. FA, EA, AFR, and LZP were responsible for visualization, formal analysis, and investigation, while RS, NS, and K contributed resources. FA, MPW, and AFR handled data curation. The original draft was prepared by FA, EA, AFR, and APAH, with all authors involved in reviewing and editing. All authors critically reviewed the manuscript for significant intellectual content and approved the final version for publication.

## Data Availability

The data underlying this study cannot be made publicly available because they contain sensitive patient-level information and are subject to ethical and confidentiality restrictions.

## ACKNOWLEDGMENTS

We thank the research assistant of the Respiratory Programmatic Implementation and Research Institute (Muhammad Ilham Dhiya Rakasiwi, MD; Fadholirrahman Raditya Naufal, MD; Vallas Aditiar, MD; Dhiya Surya Tarina, MD; Paulus Sembiring, MD; Anandya Naufal Rahadi, MD; Ahmad Fadhil Ilham, MD; Hanna Lianti Afladhia, MD; Afdila Raihani, MD; Adlina Briliani, MD; Amirah Nisrina, MD; Aulia Rizkia, MD; Ginindha Izzati S, MD; Nadya Aliza Mulyadi, MD; Rizkha MD; Gabrielle Adani, MD; Irene Audrey, MD; Nayaka Nayottama Pamadi, BPH) for their contribution to data collection and transcription.

## SUPPLEMENTARY TABLE AND FIGURE

**Supplementary Figure 1.**
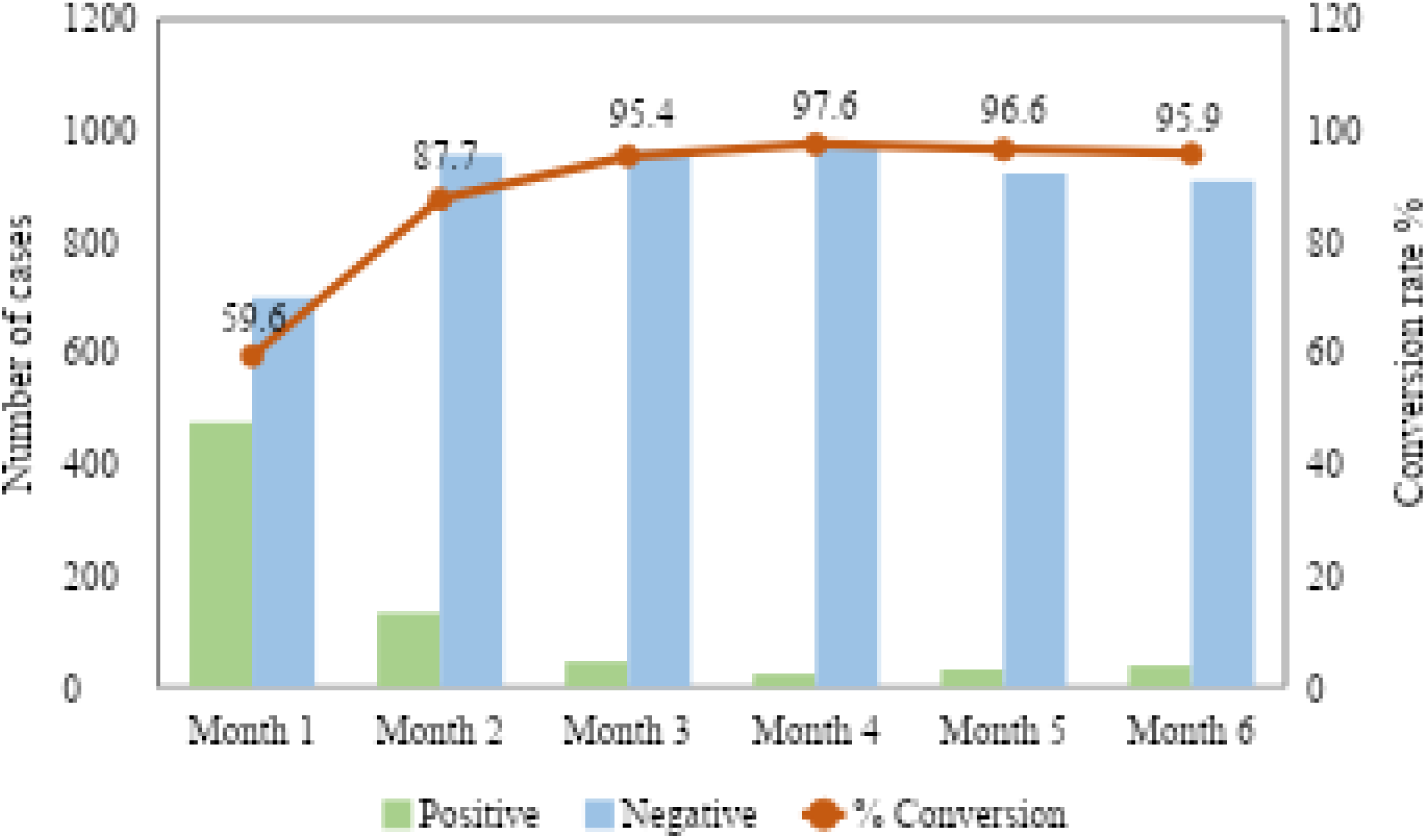
Monthly Culture Conversion Results for Monitoring DR-TB Treatment.

**Supplementary Table 1.**
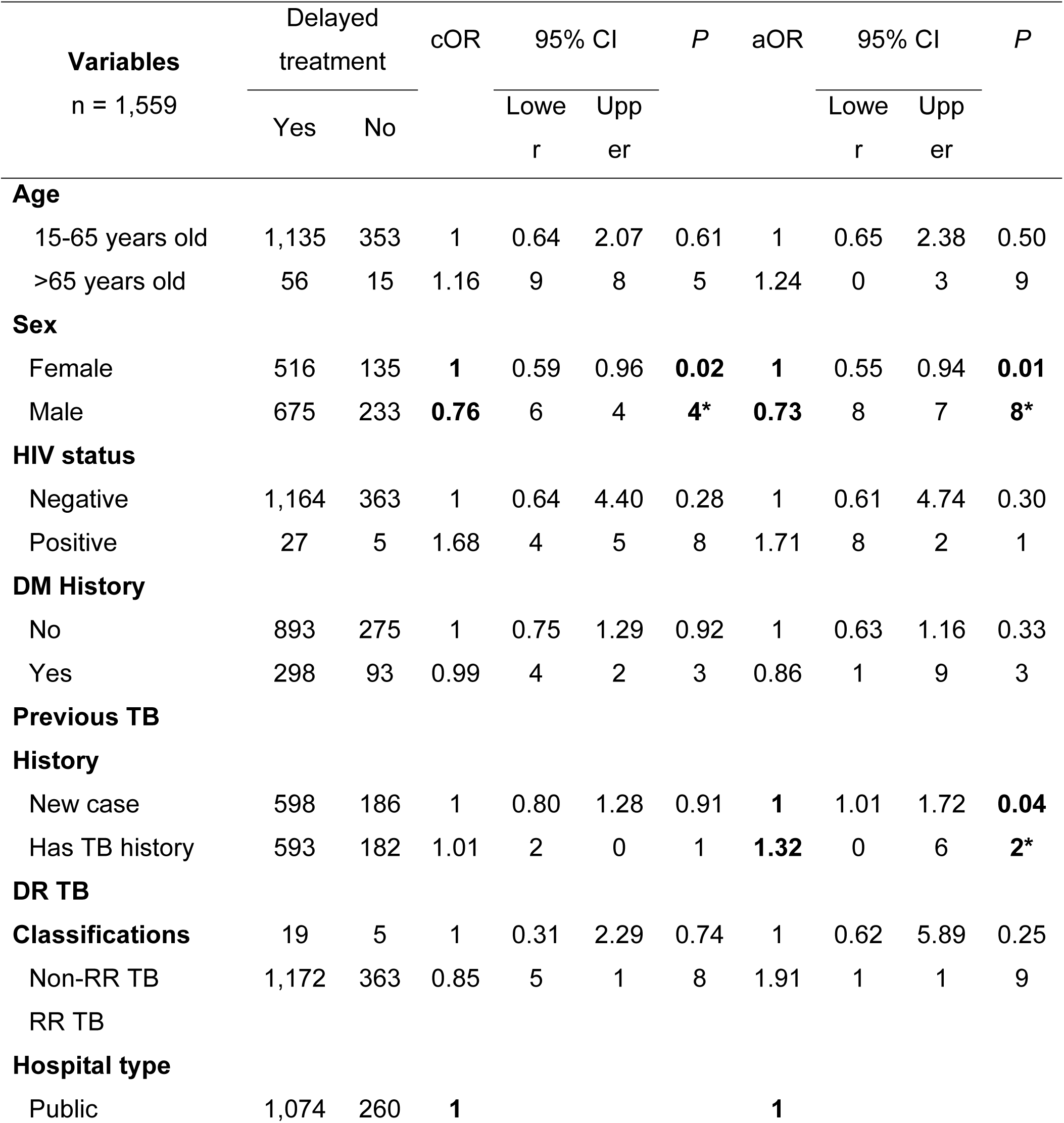
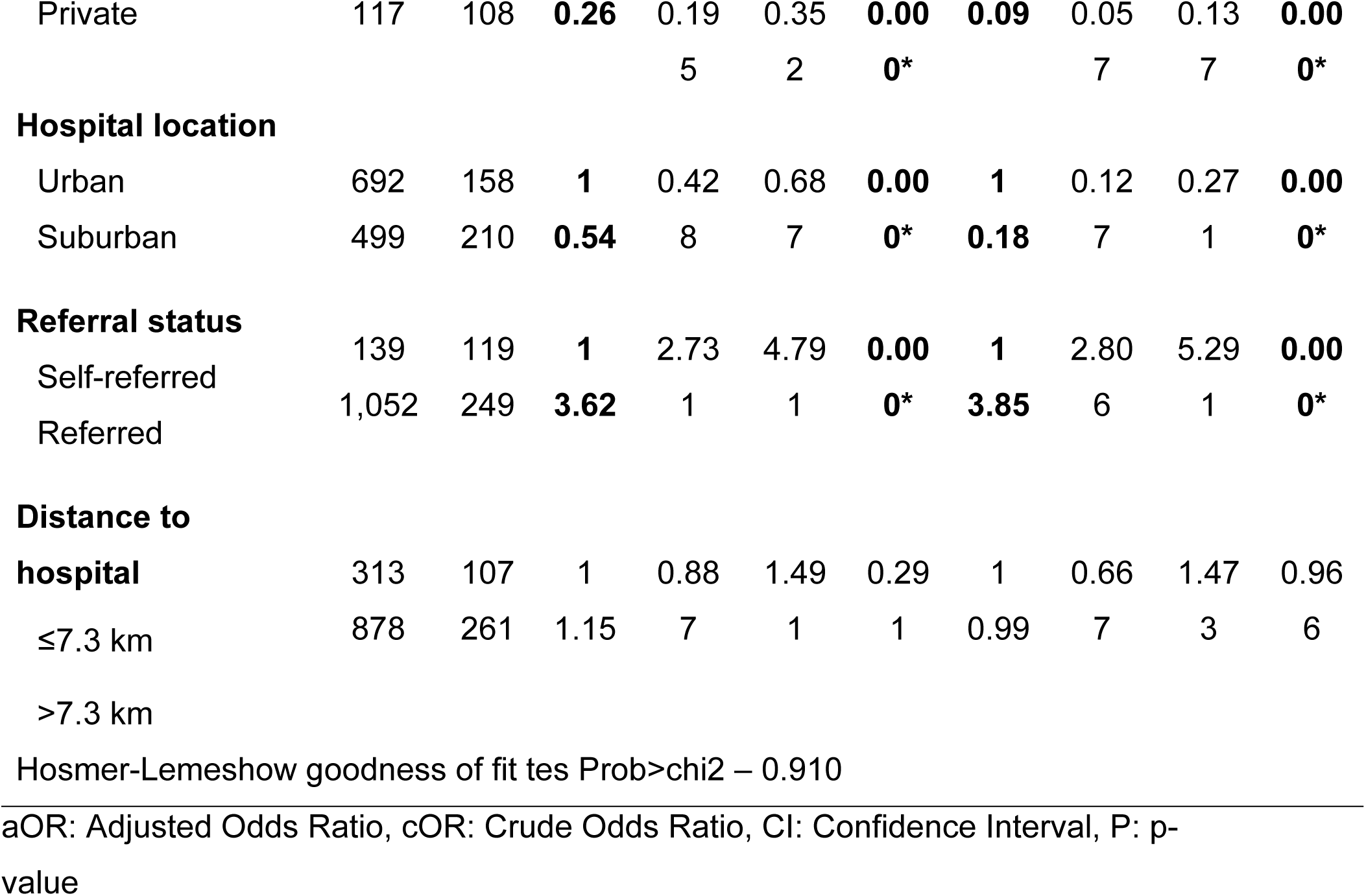
Univariable and Multivariable Logistic Regression of Factors Associated with Treatment Delay.

**Supplementary Table 2.** Univariable Logistic Regression of Treatment Interval and Culture Conversion.

| Variables<br>(Ref : negative) | Culture result |  | OR | 95% CI |  | P |
| --- | --- | --- | --- | --- | --- | --- |
|  | Positive | Negative |  | Lower | Upper |  |
| Month 1 (n=1,177) | 476 | 701 | 0.99 | 0.993 | 1.002 | 0.291 |
| Month 2 (n=1,090) | 134 | 956 | 1.00 | 0.993 | 1.008 | 0.944 |
| Month 3 (n=1,010) | 46 | 964 | **1.01** | 1.001 | 1.018 | **0.023**<br>* |
| Month 4 (n=997) | 24 | 973 | **1.01** | 1.001 | 1.021 | **0.032**<br>* |
| Month 5 (n=955) | 32 | 923 | **1.01** | 1.004 | 1.022 | **0.005**<br>* |
| Month 6 (n=949) | 39 | 910 | **1.02** | 1.006 | 1.023 | **0.000**<br>* |

OR: Odds Ratio, CI: Confidence Interval, P: p-value

**Supplementary Table 3.** Multivariate Cox Regression Analysis of Mortality in Patients with Drug-Resistant Tuberculosis.

| Variables<br>n = 1,351 | Outcome |  | HR | 95% CI |  | P |
| --- | --- | --- | --- | --- | --- | --- |
|  | mortality<br>(event) | Other<br>outcomes<br>(censored) |  | Lower | Upper |  |
| **Interval to begin treatment** |  |  |  |  |  |  |
|  | 65 | 243 | 1 | 0.69 | 1.25 | 0.665 |
| ≤7 days | 185 | 858 | 0.94 | 7 | 8 |  |
| >7 days |  |  |  |  |  |  |
| **Age** |  |  |  |  |  |  |
| 15-65 years old | 220 | 1,067 | **1** | 2.37 | 5.13 | **0.000** |
| >65 years old | 30 | 34 | **3.49** | 1 | 4 | * |
| **Sex** |  |  |  |  |  |  |
| Female | 106 | 474 | 1 | 0.72 | 1.21 | 0.633 |
| Male | 144 | 627 | 0.94 | 9 | 1 |  |
| **Previous TB History** |  |  |  |  |  |  |
| New case | 111 | 570 | 1 | 0.89 | 1.49 | 0.258 |
| Has TB history | 139 | 531 | 1.15 | 7 | 4 |  |
| **Referral status** |  |  |  |  |  |  |
| Self-referred | 53 | 167 | **1.43** | 1.03 | 1.97 | **0.029** |
| Referred | 197 | 934 | **1** | 8 | 5 | * |
| **Distance to hospital** |  |  |  |  |  |  |
| ≤7.3 km | 49 | 295 | **1** | 1.06 | 2.00 | **0.018** |
| >7.3 km | 201 | 806 | **1.46** | 8 | 9 | * |

HR: Hazard Ratio, CI: Confidence Interval, P: p-value

**Supplementary Table 4.**
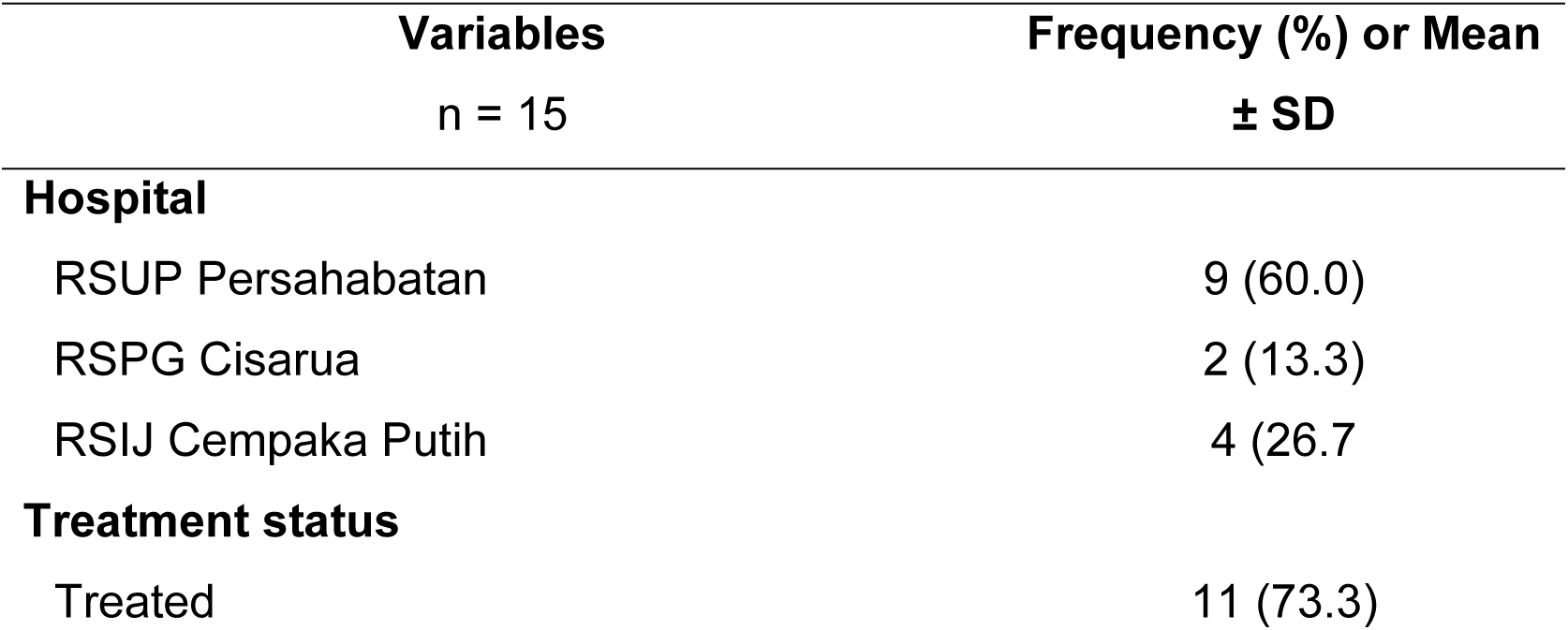
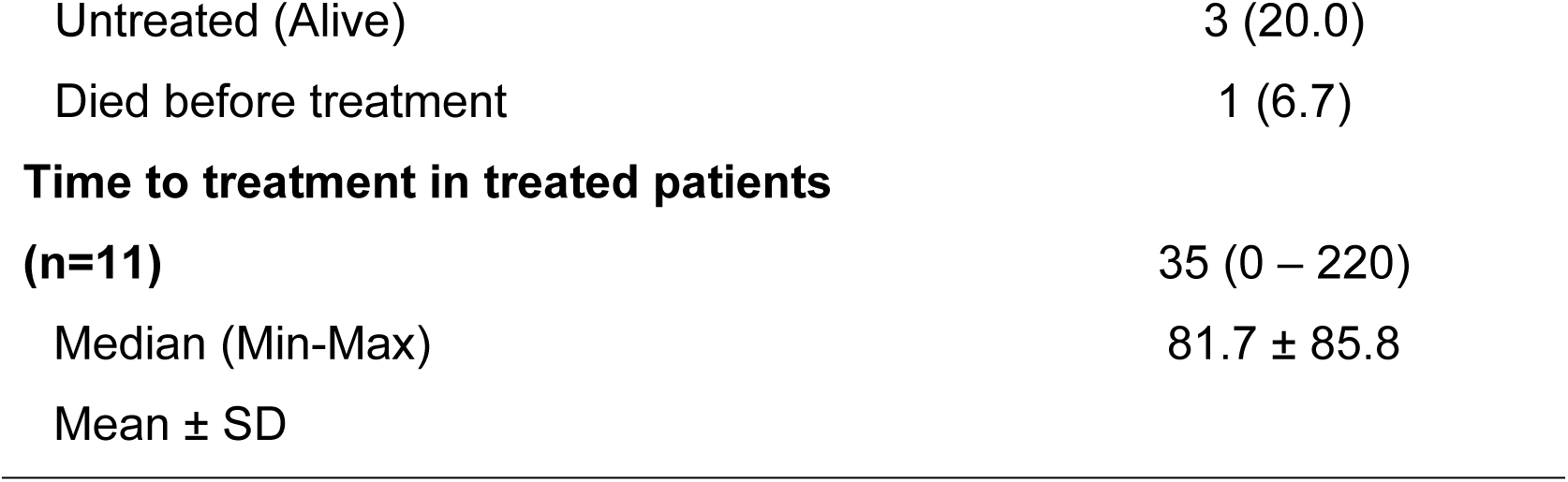
Characteristics of Informants Participating in In-depth Interview Sessions.

**Supplementary Table 5.**
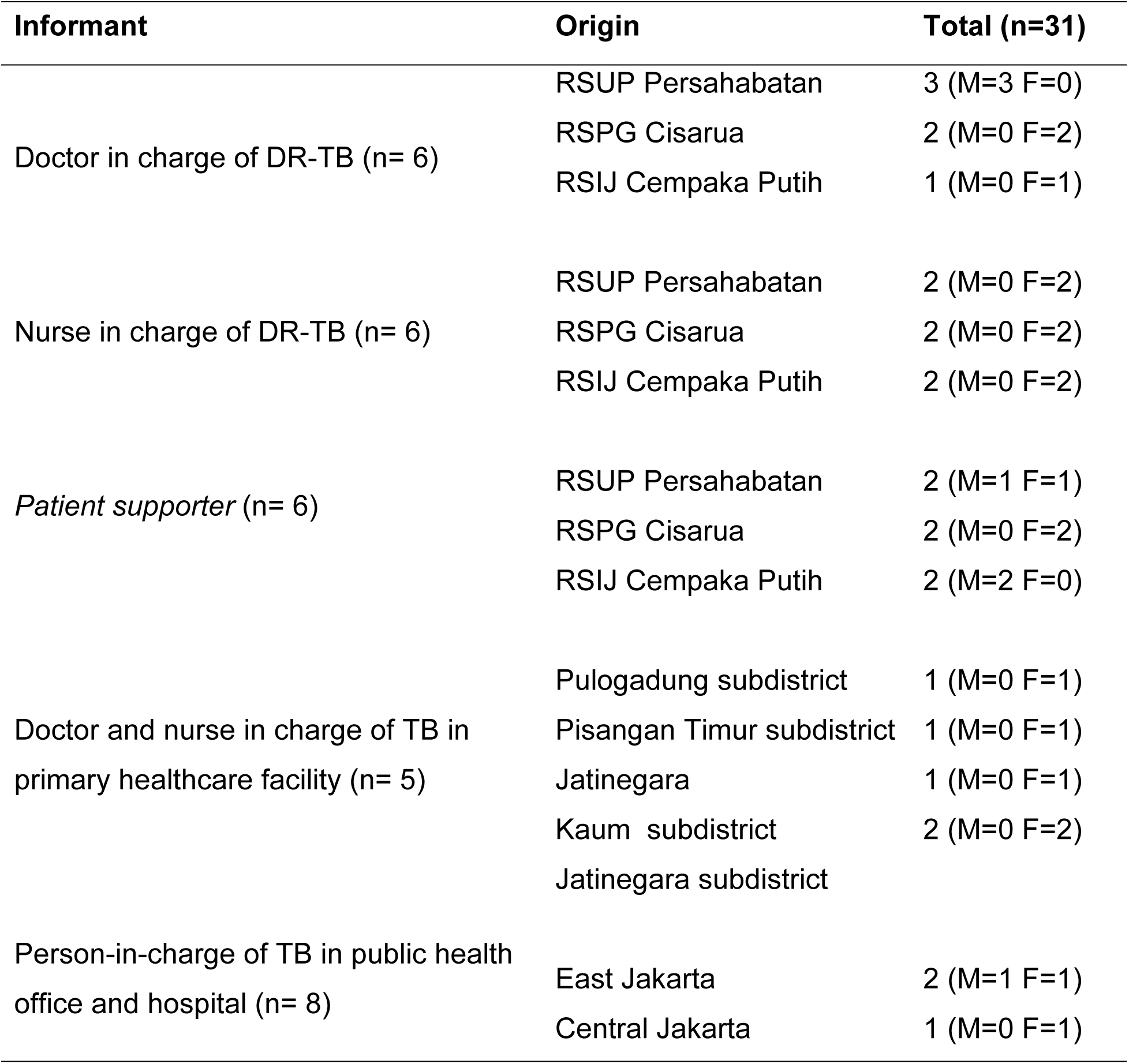
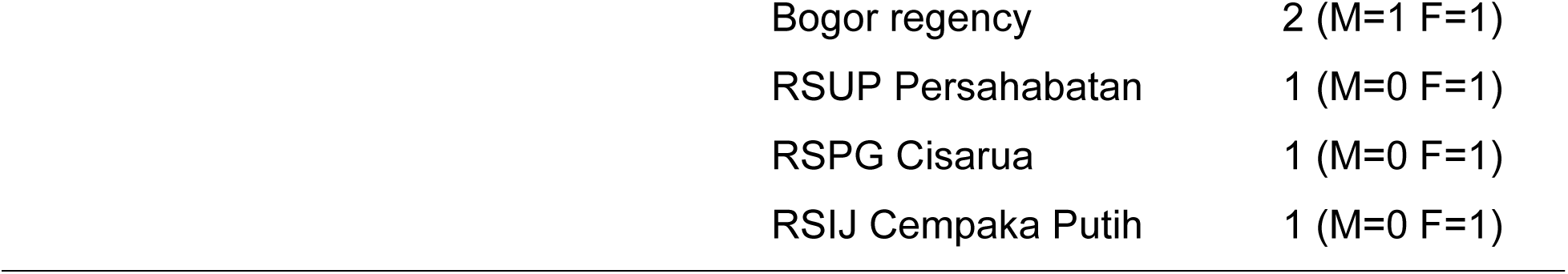
Informants Participating in Focus Group Discussion.

| Informant | Origin | Total (n=31) |
| --- | --- | --- |
| Doctor in charge of DR-TB (n= 6) | RSUP Persahabatan | 3 (M=3 F=0) |
|  | RSPG Cisarua | 2 (M=0 F=2) |
|  | RSIJ Cempaka Putih | 1 (M=0 F=1) |
| Nurse in charge of DR-TB (n= 6) | RSUP Persahabatan | 2 (M=0 F=2) |
|  | RSPG Cisarua | 2 (M=0 F=2) |
|  | RSIJ Cempaka Putih | 2 (M=0 F=2) |
| *Patient supporter* (n= 6) | RSUP Persahabatan | 2 (M=1 F=1) |
|  | RSPG Cisarua | 2 (M=0 F=2) |
|  | RSIJ Cempaka Putih | 2 (M=2 F=0) |
| Doctor and nurse in charge of TB in primary healthcare facility (n= 5) | Pulogadung subdistrict | 1 (M=0 F=1) |
|  | Pisangan Timur subdistrict | 1 (M=0 F=1) |
|  | Jatinegara | 1 (M=0 F=1) |
|  | Kaum subdistrict | 2 (M=0 F=2) |
|  | Jatinegara subdistrict |  |
| Person-in-charge of TB in public health office and hospital (n= 8) | East Jakarta | 2 (M=1 F=1) |
|  | Central Jakarta | 1 (M=0 F=1) |

|  |  |
| --- | --- |
| Bogor regency | 2 (M=1 F=1) |
| RSUP Persahabatan | 1 (M=0 F=1) |
| RSPG Cisarua | 1 (M=0 F=1) |
| RSIJ Cempaka Putih | 1 (M=0 F=1) |

**Supplementary table 6.**
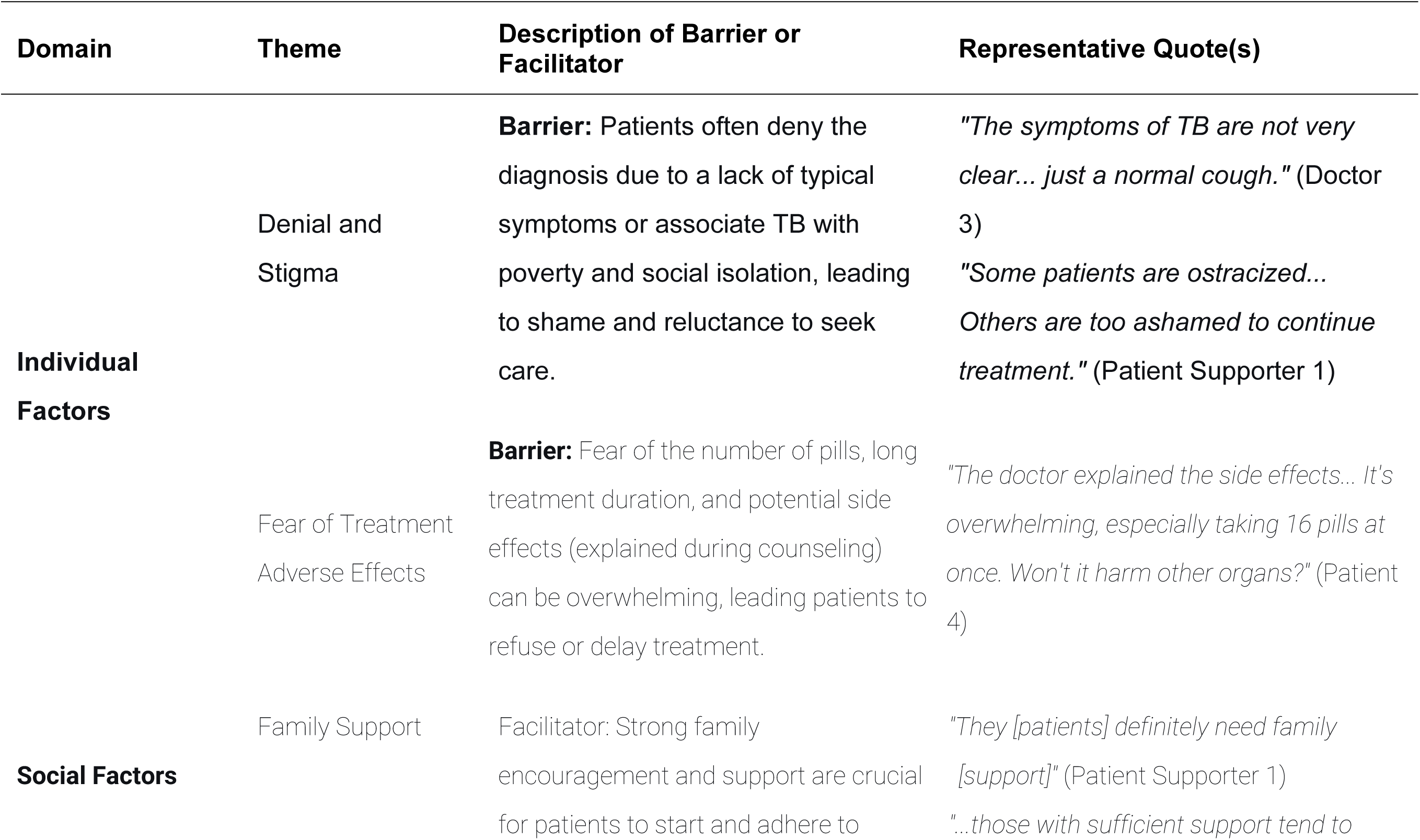
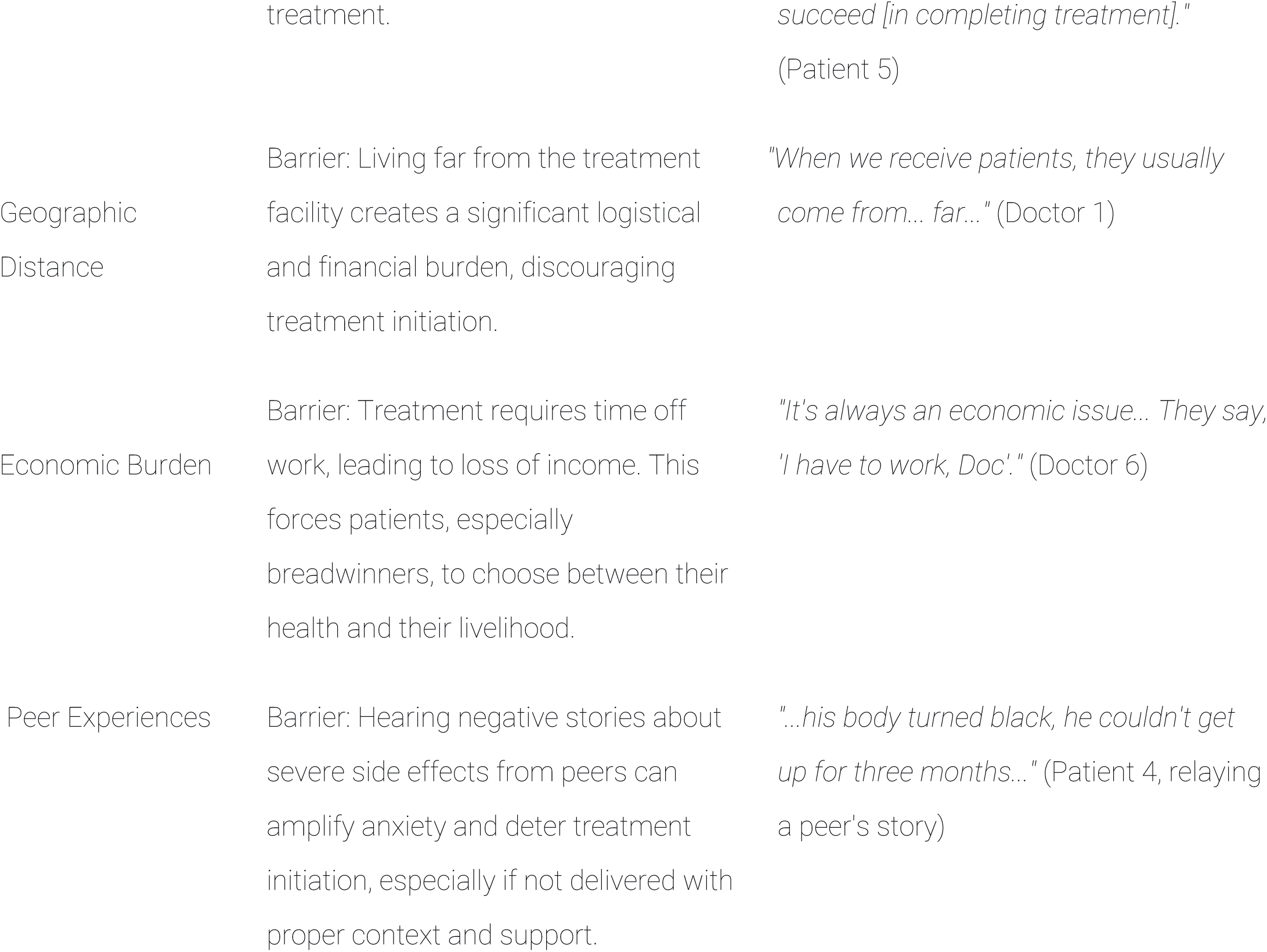
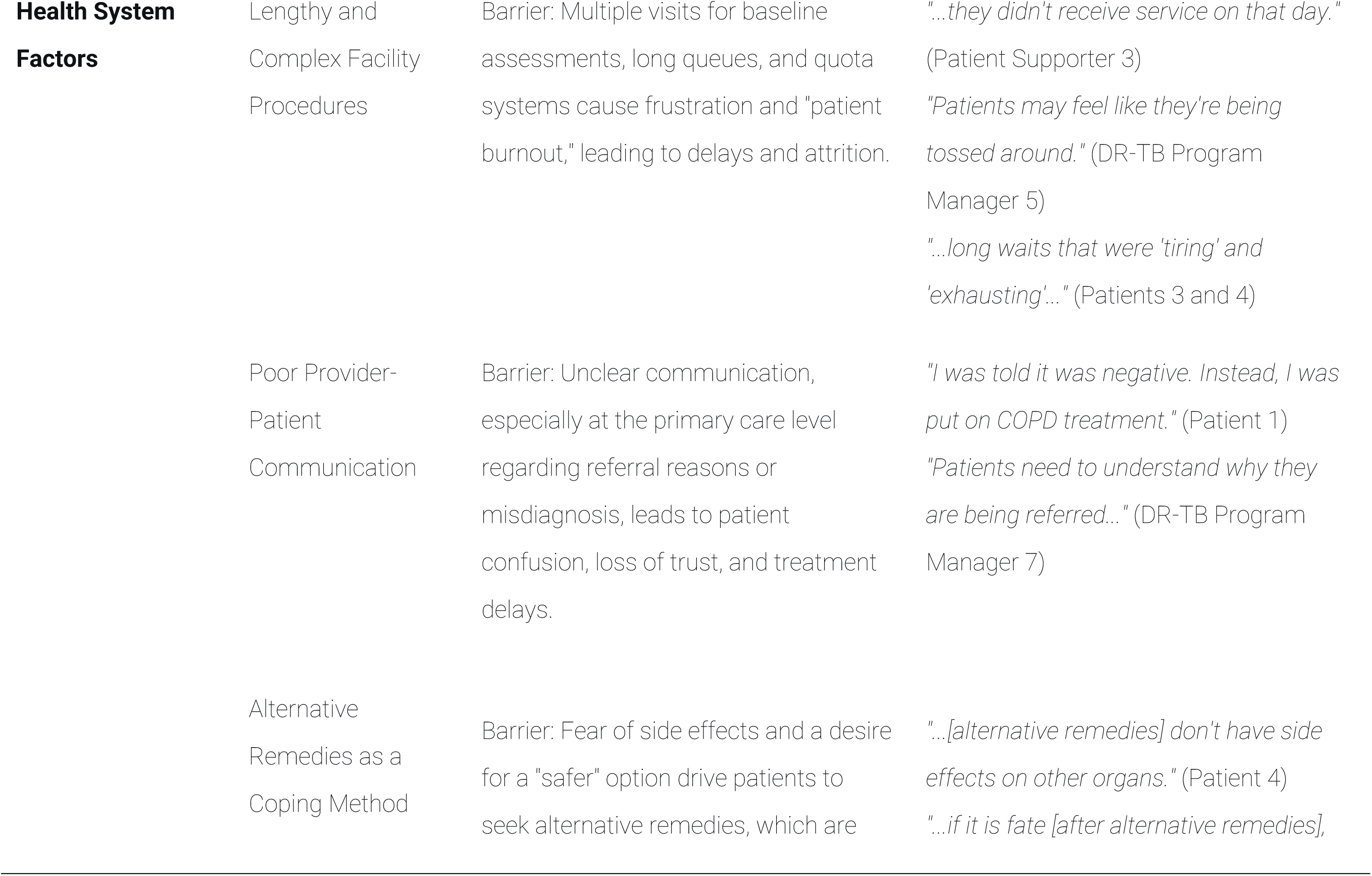
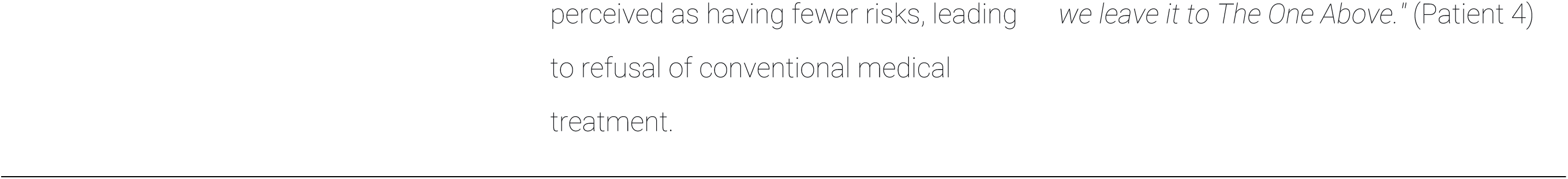
Qualitative analysis results table with representative quotations.

## Notes

### Competing Interest Statement

The authors have declared no competing interest.

### Author Declarations

The study received ethical approval from the Ethics Committee of the National Respiratory Center, RSUP Persahabatan (Approval Number: 0172.A.1/KEPK-RSUPP/12/2024, dated December 27, 2024).

